# MAGIC Composite Score Predicts Outcomes of Second-Line Therapy for Acute GVHD

**DOI:** 10.64898/2026.07.09.26357664

**Authors:** Tara Sebastian, Daniela Weber, Aaron M. Etra, Ingrid Vasova, Francis Ayuk, Hannah K. Choe, Zachariah DeFilipp, Francesco Quagliarella, Karni Bedirian, Márcio Augusto Diniz, Paibel Aguayo-Hiraldo, Peter Bader, Janna Baez, Chantiya Chanswangphuwana, Gilbert Eng, Thomas Francke, Elizabeth O. Hexner, Nikolaos Katsivelos, Carrie L. Kitko, Sabrina Kraus, Ioannis E. Louloudis, George Morales, Ryotaro Nakamura, Timothy S. Olson, Muna Qayed, Pavan Reddy, Ran Reshef, Tal Schechter, Tingyu Wang, Matthias Wölfl, Rachel Young, Robert Zeiser, William J. Hogan, John E. Levine, James L.M Ferrara

**Author notes:** **Corresponding Authors:** James L.M Ferrara, MD, DSc and John E. Levine, MD, MS, The Tisch Cancer Institute, Icahn School of Medicine at Mount Sinai, 1 Gustave L. Levy, Place/Box 1410, New York, NY 10029, Phone: (general office). W.J.H., J.E.L., and J.L.M.F. contributed equally to this work. **Data Sharing Statement:** For original data, please contact the corresponding authors listed above.

## Abstract

Approximately 30% of patients with acute graft-versus-host disease (GVHD) develop steroid-refractory disease and have very poor outcomes. Ruxolitinib has become the standard of care for steroid-refractory acute GVHD, but it is unclear which patients derive benefit. The MAGIC Composite Score (MCS), an algorithm that combines clinical symptoms and biomarkers, has been validated to predict outcomes at the start of primary GVHD treatment. Here, we evaluated its performance at the initiation of second-line treatment in 278 patients. MCS stratified patients into three risk groups (MCS1-3), with the majority (88%) classified as intermediate or high risk. Increasing MCS score was associated with progressively higher 1-year non-relapse mortality (NRM) rates (16%, 41%, and 73%; p<0.001), lower 1-year survival (77%, 56%, and 24%; p<0.001), and lower complete response (CR) rates at day 28 (47%, 38%, and 20%, respectively; p<0.01). The area under the receiver operating characteristic curve (AUROC) for 1-year NRM was significantly higher with MCS compared to clinical symptoms alone (0.70 vs. 0.63; p=0.023). Among patients treated with ruxolitinib, higher MCS similarly predicted higher NRM and lower survival and CR rates. Patients classified as MCS2/3 had poor outcomes despite ruxolitinib, underscoring the need for novel therapies in this patient population. In conclusion the MCS is an accurate predictor of outcomes for patients who require second-line treatment and may be of use as an eligibility criterion for future clinical trials in this high-risk population.

**Key Points:**

1. MAGIC Composite Scores that integrate clinical symptoms and biomarker scores predict outcomes of second-line therapy for acute GVHD more accurately than clinical symptoms alone.
2. MCS identifies patients with high NRM despite treatment with ruxolitinib.

## INTRODUCTION

Acute graft-versus-host disease (GVHD) occurs in 30-40% of patients receiving hematopoietic cell transplantation (HCT) and continues to be the leading cause of early non-relapse mortality (NRM)^1–3^. Approximately 30% of patients who receive treatment for acute GVHD develop steroid-refractory disease and require additional lines of therapy^4–7^. Outcomes for steroid-refractory patients are poor^5–10^. Ruxolitinib, an oral Janus Kinase (JAK) 1/2 inhibitor, was approved in 2019 for second-line treatment following a successful phase 2 trial, and a phase 3 randomized controlled trial confirmed its effectiveness^10–12^. Although ruxolitinib is now widely used as a standard of care second-line agent for acute GVHD, long-term outcomes with ruxolitinib demonstrate that challenges remain, especially for those patients with severe disease^13^.

The Minnesota Risk system uses the severity of clinical symptoms at acute GVHD onset to stratify patients for NRM and treatment response, but it does not identify a low-risk category suitable for treatment de-escalation^14,15^ and has not been validated for use at the start of second-line therapy. Over the last two decades, serum biomarkers of acute GVHD have consistently been able to stratify risk^16–19^. Two Mount Sinai Acute GVHD International Consortium (MAGIC) biomarkers, suppressor of tumorigenesis 2 (ST2) and regenerating islet-derived 3α (REG3α), are combined to form a single value, the MAGIC Algorithm Probability (MAP)^19^. The MAP is considered a liquid biopsy of GVHD damage to intestinal crypts^20^ and predicts mortality and treatment response at several critical timepoints, including steroid-resistant GVHD^5,19,20^.

Recently, an integrated MAGIC Composite Score (MCS) that combines clinical symptoms and biomarkers at the start of acute GVHD treatment has been shown to predict first line outcomes better than clinical symptoms alone^4^. At GVHD onset, clinical symptoms and biomarker levels interact so that NRM for a given clinical risk increases with higher biomarker scores and NRM for a given biomarker score increases with worsening clinical symptoms. In this study, we evaluated patients receiving second-line therapy for steroid-resistant GVHD at its initiation to determine whether the MCS that integrates clinical symptoms and biomarkers would better predict long-term outcomes than clinical symptoms alone.

## METHODS

### Patient Selection

We studied patients in the MAGIC database and biorepository between January 2016 and December 2025 who received second-line treatment for grade II-IV acute GVHD after initial treatment that included systemic steroids. The MAGIC database collects from 23 HCT centers in North America, Europe, and Asia using a rigorous prospective-specimen-collection, retrospective-blinded-evaluation (PRoBE) study design^22^. Informed consent was obtained from all participants in accordance with the Declaration of Helsinki under Institutional Review Board (or local equivalent) approved protocols at each center.

### Clinical GVHD Data

The clinical severity of acute GVHD was staged according to published guidelines^23^. Day 28 treatment response was assessed from the initiation of second-line treatment. Complete response (CR) was defined as complete resolution of GVHD symptoms in all target organs. Very good partial response (VGPR) was defined as only skin stage 1 symptoms remaining. Partial response (PR) was defined as improvement in at least 1 organ without worsening in other organs. All other responses, initiation of third-line treatment, and/or death before day 28 were considered non-response (NR).

### Serum Samples

Serial serum samples were collected, cryopreserved, and shipped to a central laboratory. Samples collected within three days of initiation of second-line therapy met the criteria for analysis. Serum concentrations of ST2 and REG3α were analyzed by enzyme-linked immunosorbent assays, as previously reported^24,25^. The MAP was calculated as a single value between 0.001 and 0.999 according to the formula: log[–log(1 – MAP)] = −11.263 + 1.844(log_10_ST2) + 0.577(log_10_REG3α)^19^. MAP was divided into three Ann Arbor score groups using previously validated thresholds (AA1 < 0.141; 0.141 ≤ AA2 < 0.291; AA3 ≥ 0.291)^4,19^.

### Statistical Analysis

The beginning of second-line treatment served as the starting point in all analyses. Primary endpoints were NRM at 12 months, probability of survival, and day 28 response. The cumulative incidence of NRM was estimated considering relapse as a competing risk and Gray’s test was used to compare groups^26^. The Kaplan-Meier method was used to estimate probability of survival and the log-rank test compared survival between groups^27^. Fisher’s exact test was used to compare response rates between groups^28^. Pairwise comparisons were adjusted using the false discovery rate method^29^. We used the area under the receiver operating characteristic curve (AUROC) and the Blanche method with marginal weighting to compare the prognostic value of the different models for NRM^30^. All statistical tests were 2-sided, and a *P* value <.05 was considered statistically significant. Statistical analyses were performed with R version 4.5.1 (R Foundation for Statistical Computing, Vienna, Austria).

## RESULTS

### Patient Characteristics

423 patients in the MAGIC database received second-line therapy after steroids for grade II-IV acute GVHD between the years 2016-2025 (Figure S1). 278 of these patients had serum samples available from the initiation of second-line therapy and there were no significant differences in NRM or survival between patients with or without samples (Table S1). Baseline characteristics of the study population are shown in Table 1. 64/278 (23%) patients had HLA mismatched donors. 184/278 (66%) patients had grade III-IV GVHD at the start of second-line treatment, and the vast majority (209/278, 75%) had lower GI involvement. As a result, many patients had elevated Ann Arbor (AA) scores at second-line including 140/278 (50%) patients with AA3. Approximately half of patients received ruxolitinib-based therapy as second-line treatment, with 111/278 (40%) receiving ruxolitinib alone in addition to steroids (Table S2). The median time from first to second line treatment was 11 days (range 1-230 days); 27/278 (10%) patients began second line treatment during the first three days of steroid therapy.

**Table 1.**
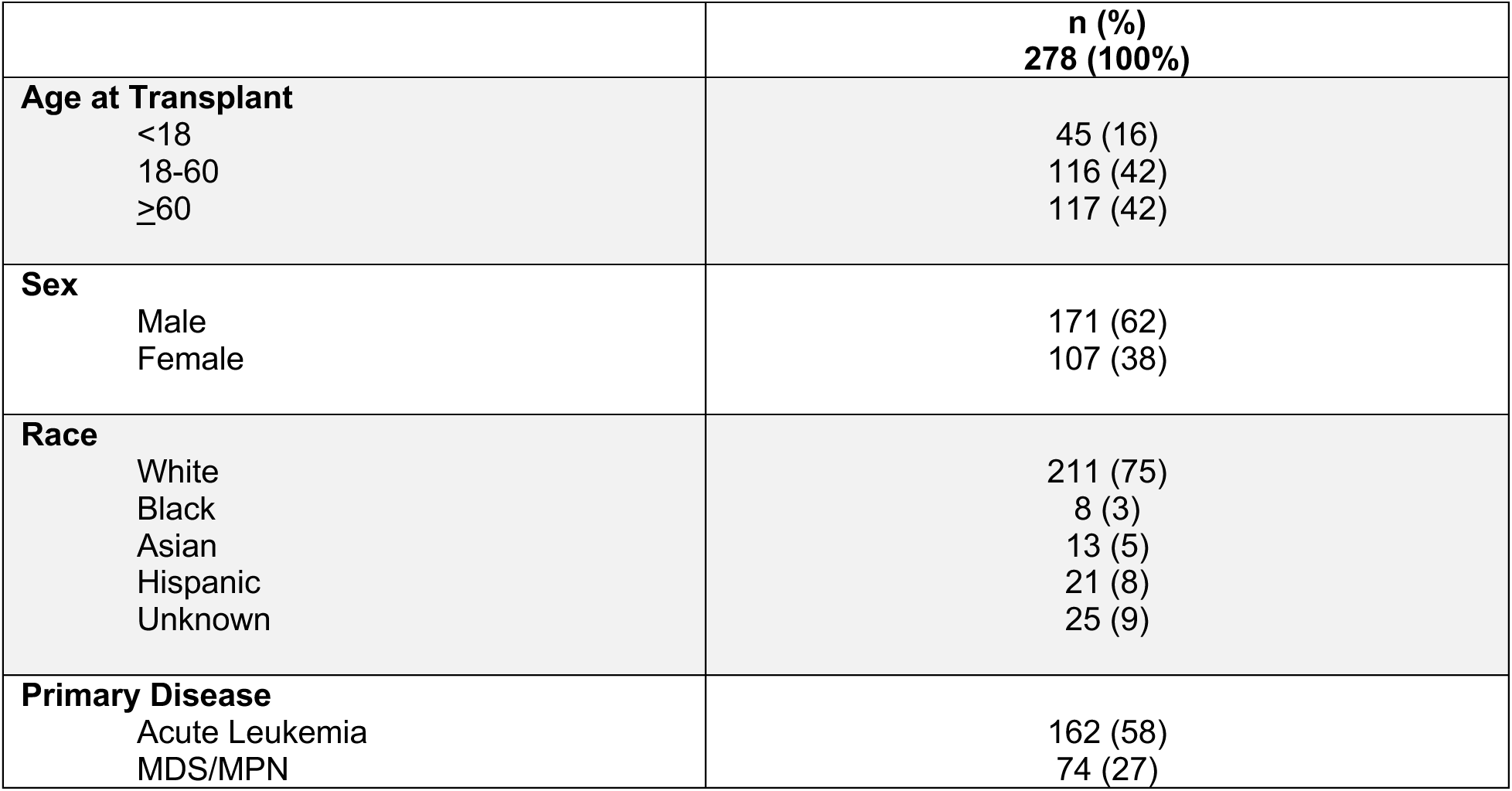
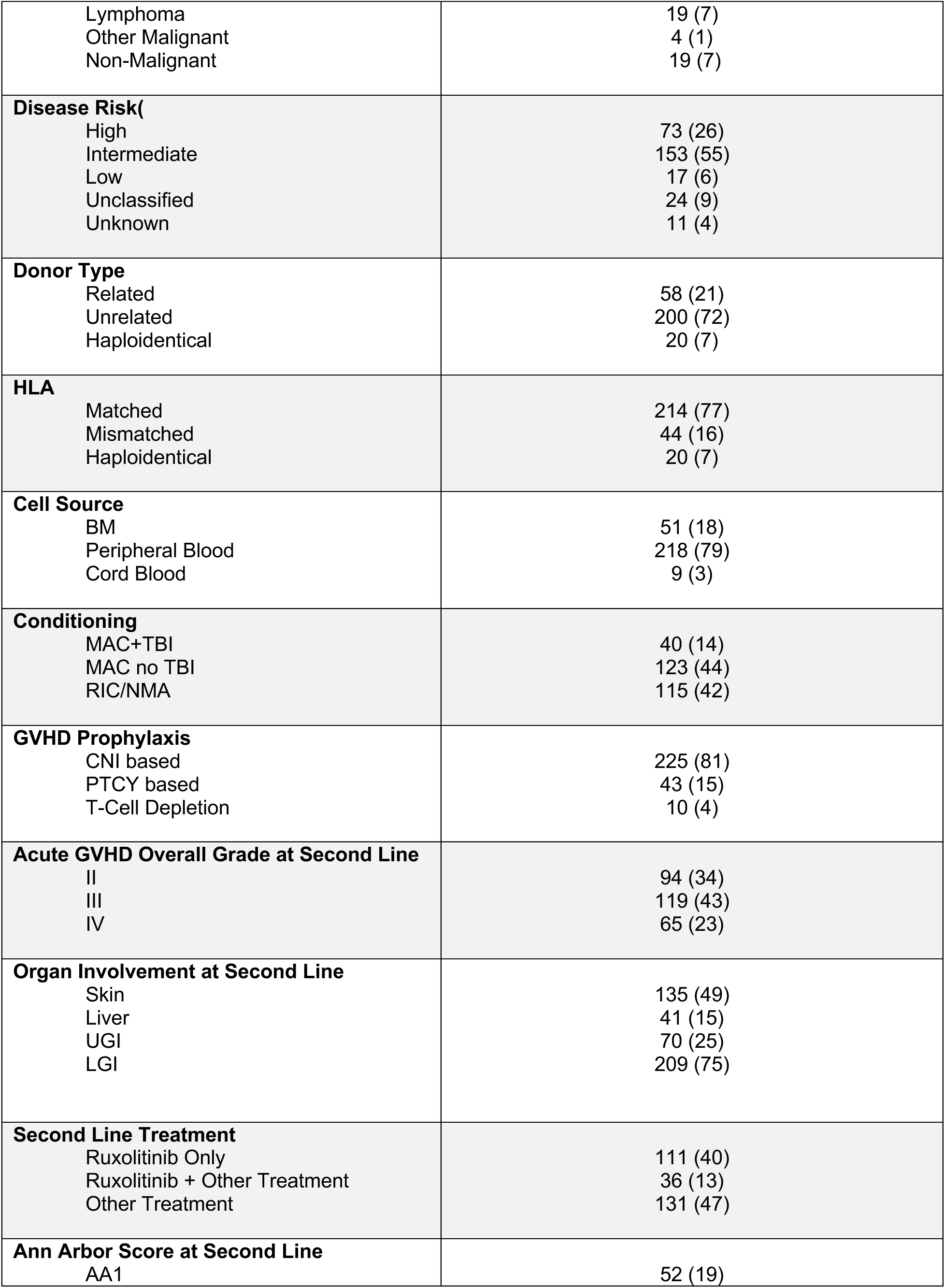
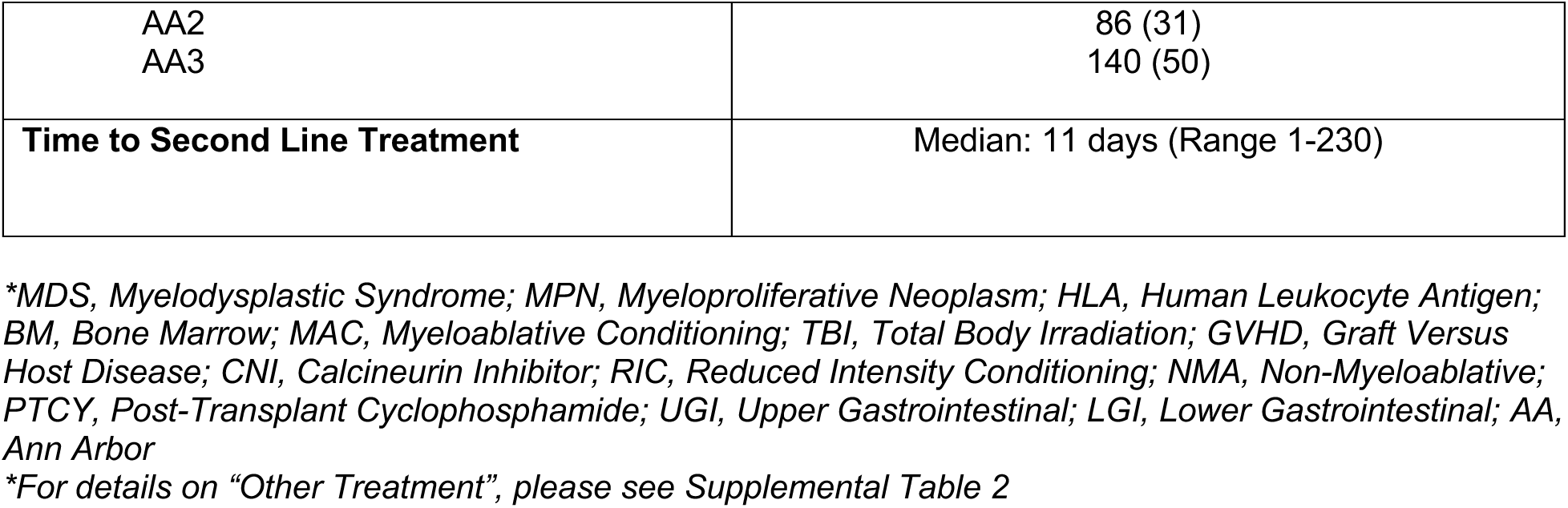
Patient Characteristics.

|  | n (%)<br>278 (100%) |
| --- | --- |
| **Age at Transplant** |  |
| <18 | 45 (16) |
| 18-60 | 116 (42) |
| ≥60 | 117 (42) |
| **Sex** |  |
| Male | 171 (62) |
| Female | 107 (38) |
| **Race** |  |
| White | 211 (75) |
| Black | 8 (3) |
| Asian | 13 (5) |
| Hispanic | 21 (8) |
| Unknown | 25 (9) |
| **Primary Disease** |  |
| Acute Leukemia | 162 (58) |
| MDS/MPN | 74 (27) |
| Lymphoma | 19 (7) |
| Other Malignant | 4 (1) |
| Non-Malignant | 19 (7) |
| **Disease Risk** |  |
| High | 73 (26) |
| Intermediate | 153 (55) |
| Low | 17 (6) |
| Unclassified | 24 (9) |
| Unknown | 11 (4) |
| **Donor Type** |  |
| Related | 58 (21) |
| Unrelated | 200 (72) |
| Haploidentical | 20 (7) |
| **HLA** |  |
| Matched | 214 (77) |
| Mismatched | 44 (16) |
| Haploidentical | 20 (7) |
| **Cell Source** |  |
| BM | 51 (18) |
| Peripheral Blood | 218 (79) |
| Cord Blood | 9 (3) |
| **Conditioning** |  |
| MAC+TBI | 40 (14) |
| MAC no TBI | 123 (44) |
| RIC/NMA | 115 (42) |
| **GVHD Prophylaxis** |  |
| CNI based | 225 (81) |
| PTCY based | 43 (15) |
| T-Cell Depletion | 10 (4) |
| **Acute GVHD Overall Grade at Second Line** |  |
| II | 94 (34) |
| III | 119 (43) |
| IV | 65 (23) |
| **Organ Involvement at Second Line** |  |
| Skin | 135 (49) |
| Liver | 41 (15) |
| UGI | 70 (25) |
| LGI | 209 (75) |
| **Second Line Treatment** |  |
| Ruxolitinib Only | 111 (40) |
| Ruxolitinib + Other Treatment | 36 (13) |
| Other Treatment | 131 (47) |
| **Ann Arbor Score at Second Line** |  |
| AA1 | 52 (19) |
| AA2<br>AA3 | 86 (31)<br>140 (50) |
| **Time to Second Line Treatment** | Median: 11 days (Range 1-230) |

\*MDS, Myelodysplastic Syndrome; MPN, Myeloproliferative Neoplasm; HLA, Human Leukocyte Antigen; BM, Bone Marrow; MAC, Myeloablative Conditioning; TBI, Total Body Irradiation; GVHD, Graft Versus Host Disease; CNL, Calcineurin Inhibitor; RIC, Reduced Intensity Conditioning; NMA, Non-Myeloablative; PTCY, Post-Transplant Cyclophosphamide; UGI, Upper Gastrointestinal; LGI, Lower Gastrointestinal; AA, Ann Arbor
\*For details on "Other Treatment", please see Supplemental Table 2

### MAGIC Composite Score Predicts Non-Relapse Mortality and Survival

We first analyzed NRM and survival by clinical grade. No significant differences in outcomes were seen between grades III and IV GVHD (Table S3), and clinical risk was, therefore, classified as either grade II or grade III/IV. Acute GVHD accounted for 77% of the deaths within 1 year of second-line therapy (Table S4). As expected, compared to patients with grade III-IV, patients with grade II GVHD demonstrated significantly less NRM (12-month NRM 28% vs. 61%, p<0.001) and better survival (12-month OS 65% vs. 37%, p<0.001) (Figure 1A).

**Figure 1.**
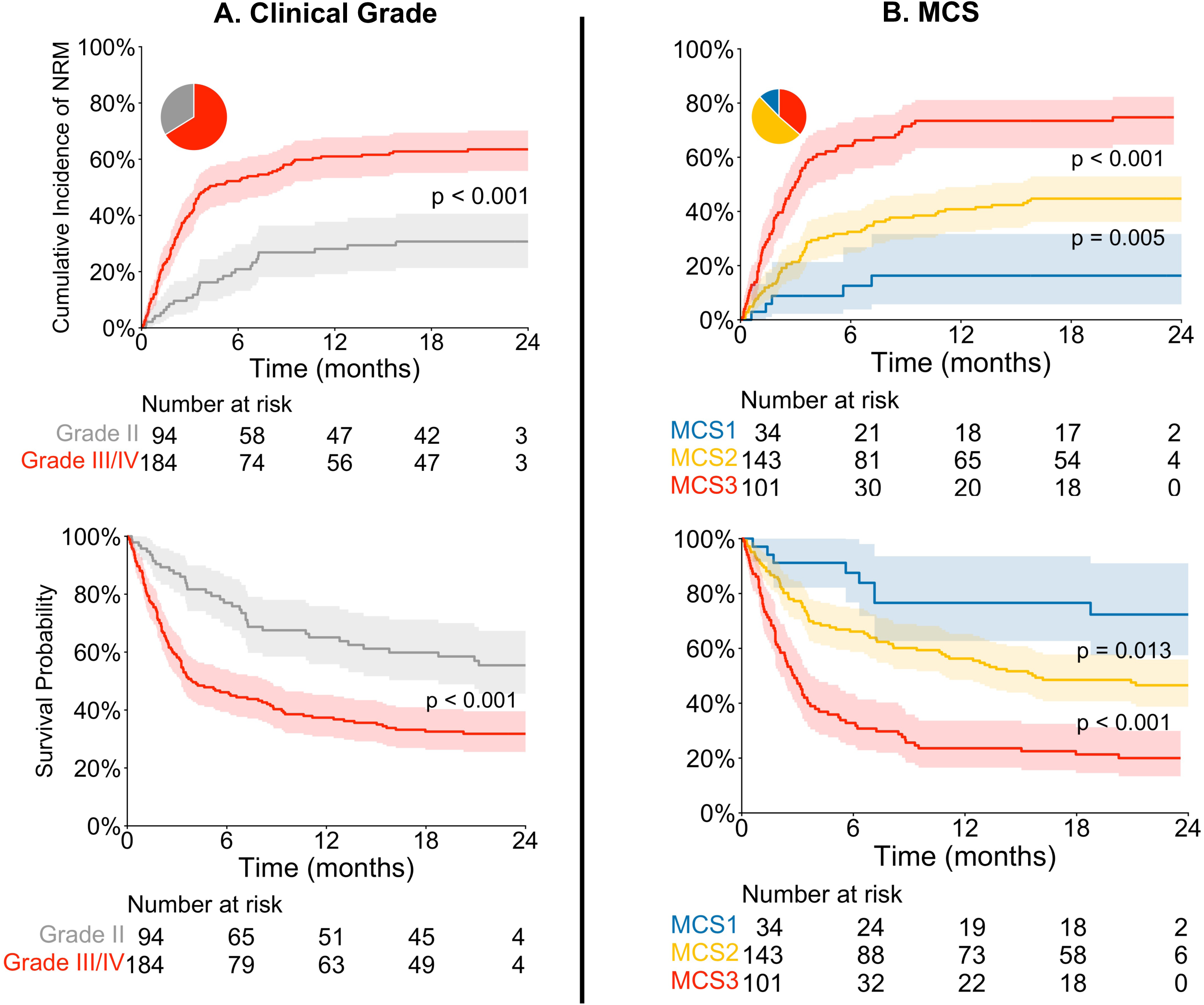
Long Term Outcomes by Risk System. Cumulative incidence of NRM and probability of survival after second-line therapy stratified by (A) clinical grade and (B) MCS. Pie charts depict the percentage of patients in each risk category.

Manhattan risk categories use clinical organ staging at treatment onset to classify patients into three separate groups. In this study, patients were first categorized based on symptoms at the start of second line into three Manhattan Risk categories as previously described^4^ (Table S5) which were then integrated with biomarker (AA) scores to create three MCS groups (Table 2). As expected, NRM increased with each AA score within each Manhattan risk group. In Manhattan intermediate risk, 12-month NRM was 16% for AA1, 30% for AA2, and 54% for AA3. NRM also increased as clinical symptoms became more severe within each AA score group. For example, in AA3 patients, 12-month NRM was 54% in Manhattan intermediate and 73% in Manhattan high groups. As a result of biomarker integration with clinical symptoms, 34/278 patients (12%) were classified as MCS1, 143/278 (51%) MCS2, and 101/278 (37%) MCS3 (Figure S2). These results demonstrate an interaction between the clinical and biomarker grading systems (Table 2).

**Table 2.** Integration of Manhattan Risk and Biomarkers into MAGIC Composite Score.

| Manhattan Risk | AA Score | 12-month NRM | MAGIC Composite Score | n (%)<br>278 (100%) |
| --- | --- | --- | --- | --- |
| Intermediate<sup>+</sup> | 1 | 16% | MCS 1 | 34 (12) |
| Intermediate<sup>++</sup> | 2 | 30% | MCS 2 | 43 (15) |
| Intermediate | 3 | 54% | MCS 2 | 38 (14) |
| High | 1 | 34% | MCS 2 | 19 (7) |
| High | 2 | 43% | MCS 2 | 43 (15) |
| High | 3 | 73% | MCS 3 | 101 (37) |

\*AA, Ann Arbor; NRM, Non-Relapse Mortality
+ Includes 2 patients with low Manhattan risk who became MCS 1
++ Includes 1 patient with low Manhattan risk who became MCS 2

This interaction between clinical and biomarker grading systems was further supported when integrated MCS scores were compared to categories by biomarkers alone. Ann Arbor scores divided patients into three risk groups, but the differences between AA1 and AA2 were not statistically significant (Figure S3). Integration of clinical symptoms reclassified a subset of low risk AA1 patients (19/52) as MCS2. This subset accounted for a substantial proportion of NRM (60%) within the AA1 group; their reclassification as intermediate risk resulted in a highly statistically significant difference in NRM between MCS1 and MCS2.

The MCS system created three distinct risk groups for NRM and survival. With each increase in MCS, the incidence of 12-month NRM significantly increased (16%, 41%, and 73%, respectively; p<0.001) (Figure 1B). No differences in relapse were seen between MCS groups (Figure S4), and thus the probability of 12-month survival significantly decreased (77%, 56%, and 24%, respectively; p<0.001) (Figure 1B). The area under the receiver operating characteristic curve (AUROC) for 12-month NRM was significantly higher in the MCS model than the clinical grade model (0.70 vs. 0.63; p=0.023). A subset analysis of patients who received second line therapy more than three days after first line therapy produced similar results (Figure S5).

### MAGIC Composite Score Stratifies Response to Second-Line Treatment

We next evaluated the day 28 response to second-line therapy. Complete response when compared to partial response was strongly associated with improved 12-month NRM (25% vs 56%, p<0.01) and was, therefore, used for these analyses (Figure S6). With each increase in MCS, day 28 CR decreased by 10-15%, although the difference between MCS1 and MCS2 did not reach statistical significance noting the small number of MCS1 patients (Figure 2). Notably, each increase in MCS also correlated with a decreased probability of day 28 overall response, the common primary endpoint for clinical trials in steroid-refractory GVHD (Figure S7). The response rates between patients categorized as grade II or MCS1 were equivalent (∼50%). The MCS classification, however, created a highly significant difference in response between intermediate (MCS2) and high-risk (MCS3) groups (p=0.006) that did not exist between clinical grades III and IV GVHD.

**Figure 2.**
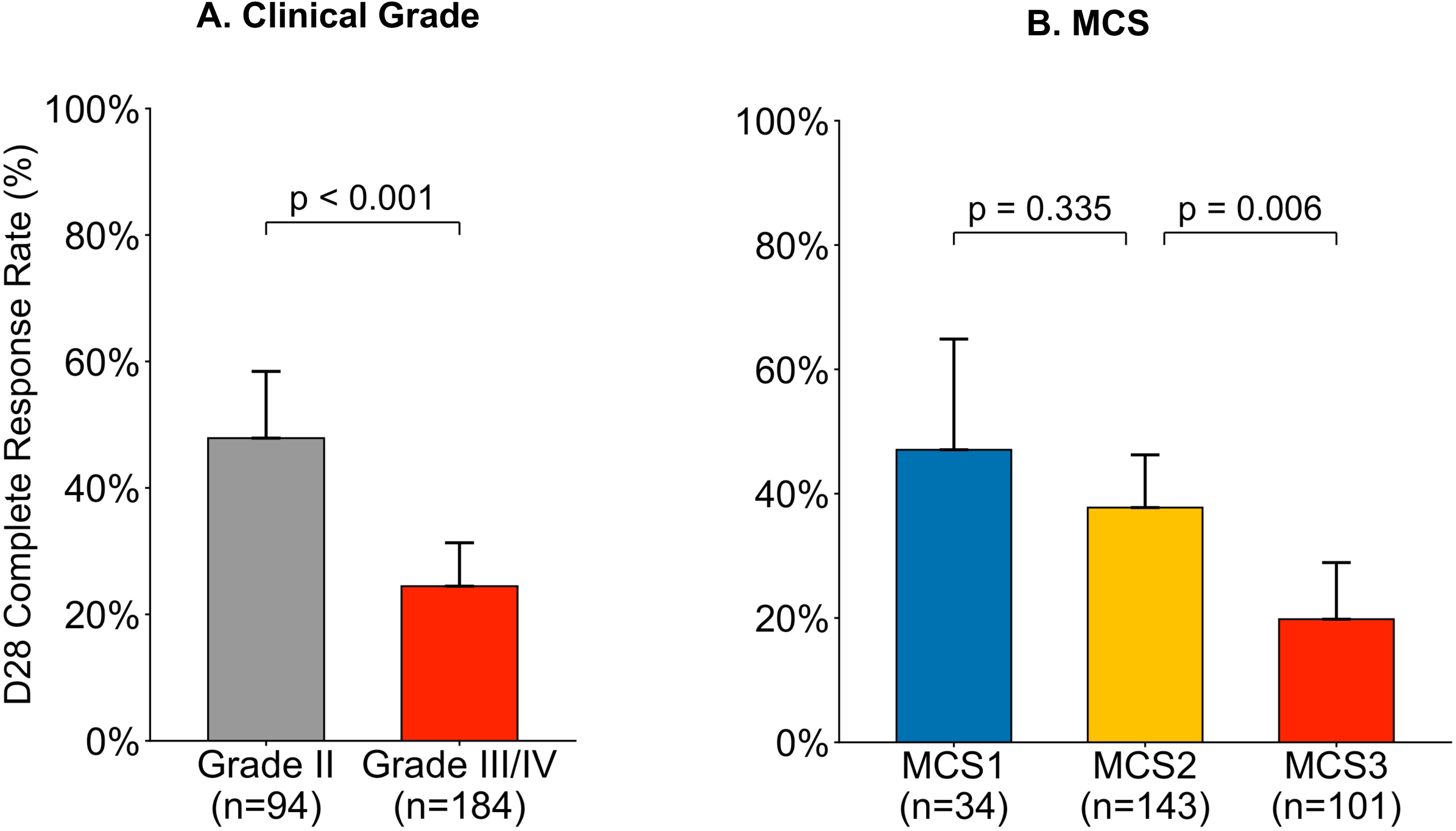
Day 28 Complete Response Rate by Risk System. D28 CR rate to second-line therapy shown according to (A) clinical grade (grade II: 50% vs. grades III/IV: 25%) and (B) MCS (MCS1: 47%, MCS2: 38%, MCS3: 20%)

### MAGIC Composite Score Predicts Outcomes After Ruxolitinib

Ruxolitinib use as a second-line therapy for acute GVHD increased dramatically following regulatory approval in 2019, from 30% of patients before to 71% thereafter (Table S6). As expected, ruxolitinib improved NRM (Figure S8) and complete response (39% vs 25%, p=0.02) compared to other second-line treatments. Due to this change in the standard of care, we analyzed our model in the subset of patients who were treated with ruxolitinib as a second-line therapy (n=147). Even with smaller numbers, MCS created three distinct groups: 12-month NRM increased (13%, 37%, and 67%, respectively; p<0.01) and D28 response rates decreased by 15-20% with each increase in MCS (Figure 3, Figure S9). Only the small subset of MCS1 patients achieved CR rates greater than 50% and corresponding low NRM. In contrast, many patients with clinical grade II GVHD and high-risk AA2/3 biomarkers were classified as MCS2 and experienced high NRM despite treatment with ruxolitinib. Small sample sizes precluded comparing ruxolitinib alone to any of the combinations of ruxolitinib with a second agent (Table S2).

**Figure 3.**
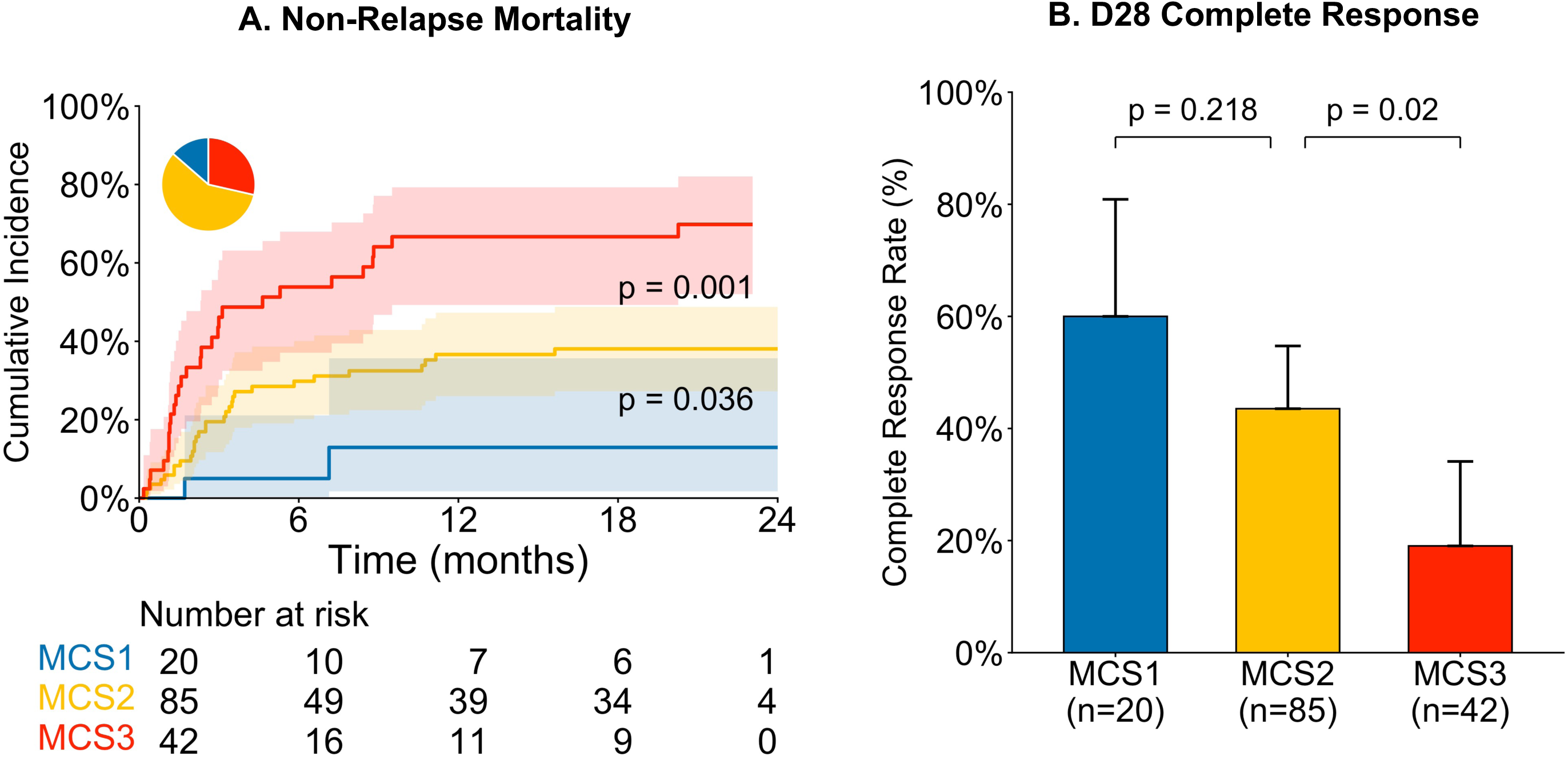
Long Term Outcomes with Ruxolitinib Treatment. Patients who received ruxolitinib for second line therapy (n= 147) were classified by MCS and analyzed for (A) cumulative incidence of NRM and (B) day 28 complete response (MCS1: 60%; MCS2: 44%; MCS3: 19%).

## DISCUSSION

In this study, we investigate the use of a novel MAGIC composite score to stratify risk at second-line therapy for steroid-refractory GVHD. Our results demonstrate that the MCS separates patients into three distinct risk groups with significantly different NRM survival, and response rates. Incorporation of biomarkers identified patients at high risk for treatment failure and death despite ruxolitinib, including the many clinical grade II patients who have high biomarker scores at the time of initiation of second-line therapy. The poor outcomes for MCS2/3 patients, who represent the majority of patients requiring second-line treatment, emphasize the persistent need for more effective treatment strategies in this setting.

A previous study has demonstrated the utility of MAPs in identifying high-risk patients at the start of second-line therapy^5^. A key finding of our work is that biomarker levels and clinical symptoms interact in these patients as they do at the start of GVHD. Patients with steroid-refractory GVHD of the same clinical grade have vastly different outcomes based on their Ann Arbor score and patients of the same Ann Arbor score have worse outcomes with more severe clinical symptoms. NRM more than triples in Manhattan intermediate risk patients as AA score increases from 1 to 3. Similarly, NRM more than doubles for AA1 patients when Manhattan risk increases from intermediate to high, underscoring that both clinical and laboratory values are useful for accurate evaluation and risk stratification at second-line initiation as at the start of first line therapy.

Most patients in our study had persistent gastrointestinal symptoms and high AA scores despite first line corticosteroids. Corticosteroids can paradoxically impair intestinal epithelial repair by reducing the proliferative ability of intestinal stem cells, a phenomenon termed the “tissue repair gap”^31–33^. MAP scores function as a liquid biopsy of the intestine^19^, such that AA2 and AA3 scores following systemic steroid therapy reflect significant GI crypt damage and a persistent tissue repair gap. Only a small subset of patients with steroid-refractory GVHD had AA1 (and corresponding MCS1) scores demonstrating adequate regenerative ability. The superior discrimination of 12-month NRM by MCS compared with day 28 clinical response (complete or overall) further supports this biological framework. A positive symptom-based response may fail to capture persistent, subclinical tissue damage that can be detected by biomarkers and that later manifests as treatment failure or death. In contrast, response criteria integrating both clinical and biomarker data more accurately reflect underlying disease biology and provide stronger surrogates for long-term outcomes^34^.

These findings suggest that most patients with MCS2/3 status after steroid treatment are unlikely to achieve good outcomes with ruxolitinib alone and will require additional therapies that promote intestinal healing and epithelial repair^33^. While ruxolitinib has shown direct activity promoting intestinal stem cell proliferation in preclinical models^33^, combination strategies such as the planned trial of ruxolitinib with mesenchymal stromal cells (BMT CTN 2303) are important avenues for investigation. More broadly, these results highlight the need for improved first line therapies that minimize corticosteroid use to prevent gastrointestinal damage.

This study has several limitations. First, the size of the study population is relatively small, particularly for patients who received second-line ruxolitinib. As in most HCT studies, some demographic groups are underrepresented, such as children, Black, and Hispanic patients. Many patients did not have samples available at the start of second-line therapy. We approached this issue by performing a sensitivity analysis and found no significant difference in NRM or OS for patients with vs without samples. Furthermore, some analyses lack the power needed to achieve statistical significance. Second, sample size was not large enough to explore interactions between MCS and other clinical risk factors such as timing and combination of second-line therapy. Third, patients with grade I disease who received second-line therapy were excluded from the study because systemic treatment is not typically recommended for this group, even though these patients may reflect real-world clinical practice. Fourth, a few patients were included who received second line treatment within the first 3 days of steroid therapy and do reflect evolving real-world practices. Their inclusion did not affect the results, but their treatment does not conform to the common criteria for steroid-refractory GVHD^10^. Patients who received ruxolitinib in combination with other therapies also reflect real-world practice but limited our ability to characterize ruxolitinib’s effect in isolation.

In conclusion, the MCS model which integrates clinical symptoms and biomarker scores creates three distinct risk groups for steroid-refractory patients at the initiation of second-line therapy that predict NRM, survival, and response to therapy. Outcomes remain very poor overall, perhaps in part due to lack of, or even impairment of, intestinal tissue repair by steroids. This model offers the potential to identify high-risk patients in need of novel second-line therapies.

## Data Availability

All data produced in the present study are available upon reasonable request to the authors.

## Acknowledgements

This work was supported by the National Cancer Institute (NCI) (P01 CA039542, P30 CA196521), National Heart, Lung, and Blood Institute (NHLBI) (R21 HL177578), the National Institute of Allergy and Infectious Disease (NIAID) (1R38AI181012-01), the Pediatric Cancer Foundation, the Solomon and Gillespie Fund, Deutsche Forschungsgemeinschaft (DFG) (324392634 - TRR 221), and Jose Carreras Leukämie Stiftung (Grant DLCS 01 GvHD/2020).

## Authorship Contributions

T.S. designed the study, collected the clinical data, performed laboratory experiments, conducted the statistical analysis, and wrote the manuscript; K.B. and M.A.D. advised statistical methods. G.M. performed laboratory experiments. I.E.L. and T.W. collected and reviewed clinical data and advised statistical methods. D.W., A.M.E, I.V., F.A., H.K.C., Z.D., F.Q., P.A.H., P.B., J.B., C.C., G.E., T.F., E.O.H., N.K., C.L.K., S.K., R.N., T.S.O., M.Q., P.R., R.R., T.Sc., M.W., R.Y., and R.Z. collected and reviewed clinical data and revised the manuscript. W.J.H, J.E.L. and J.L.M.F. designed the study, interpreted data, advised methods, reviewed, and revised the manuscript, and organized this project. All authors contributed to the writing of the report and approved the final version of the article.

