## Supplemental Materials for "MAGIC Composite Score Predicts Outcomes of Second-Line Therapy for Acute GVHD"

**Table S1.** Survival and NRM for Patients With and Without Samples

| | With Samples (95% CI) n=278 | Without Samples (95% CI) n=145 | p-value |
|---|---|---|---|
| **12-month Survival** | 47% (41-53) | 54% (46-63) | 0.25 |
| **12-month NRM** | 50% (44-56) | 38% (30-46) | 0.1 |

**Table S2.** Treatments Used for Second Line Therapy

| | 2nd Line Therapy | n |
|---|---|---|
| **Ruxolitinib Monotherapy** | Ruxolitinib | 111 |
| **Ruxolitinib + Other Therapies+** | ECP | 11 |
| | Vedolizumab | 7 |
| | Tocilizumab | 5 |
| | Basiliximab | 4 |
| | Etanercept | 4 |
| | Infliximab | 3 |
| | Rituximab | 2 |
| | AAT | 2 |
| | MMF | 2 |
| | MSC | 1 |
| | Abatacept | 1 |
| **Other Therapies++ (Without Ruxolitinib)** | ECP | 38 |
| | Etanercept | 34 |
| | Vedolizumab | 21 |
| | Tocilizumab | 18 |
| | MMF | 14 |
| | ATG/Alemtuzumab | 11 |
| | AAT | 6 |
| | Basiliximab | 6 |
| | MSC | 5 |

| | | |
|---|---|---|
| | Tacrolimus | 5 |
| | Infliximab | 3 |
| | Rituximab | 2 |
| | UVA | 2 |
| | Brentuximab | 1 |
| | CSA | 1 |
| | Everolimus | 1 |
| | Fecal Microbial Transplant | 1 |
| | Itacitinib | 1 |
| | Natalizumab | 1 |
| | Sirolimus | 1 |
| | Anti-CD3/CD7 immunotoxin | 1 |

**ECP, Extracorporeal Photopheresis; AAT, Alpha -1 Antitrypsin; MMF, Mycophenolate Mofetil; MSC, Mesenchymal Stromal Cells; ATG, Anti-Thymocyte Globulin; UVA, Ultraviolet A; CSA, Cyclosporine A*
*+Includes 6 patients who received 3 therapies and are counted in multiple categories*
*++Includes 36 patients who received a combination of therapies and are counted in multiple categories*

**Table S3.** Outcomes Stratified by Clinical Grade

| | **Grade II (95% CI) n=94** | **Grade III (95% CI) n=119** | **Grade IV (95% CI) n=65** | **p-value** |
|---|---|---|---|---|
| **12-mo Survival** | 65% (56-76) | 41% (33-51) | 31% (22-45) | Grade II vs III: p<0.001<br>Grade III vs IV: 0.346 |
| **12-mo NRM** | 28% (20-38) | 57% (47-65) | 69% (56-79) | Grade II vs III: p <0.001<br>Grade III vs IV: p=0.161 |
| **Day 28 CR** | 48% (38-58) | 29% (21-38) | 17% (9-8) | Grade II vs III: p=0.009<br>Grade III vs IV: 0.106 |

**Table S4.** Causes of Death Within 12 Months of Second Line Therapy

| Cause of Death | n (%)<br>143 (100) |
|---|---|
| **GVHD Related** | 116 (81) |
| Acute* | 110 (77) |
| Chronic | 6 (4) |
| **Relapse** | 10 (7) |
| **Infection** (after resolution of GVHD) | 7 (5) |
| **Other** | 7 (5) |
| Cardiac Event | 2 (1) |
| Liver/Renal Toxicity | 1 (0.7) |
| Post-Transplant Lymphoproliferative Disease | 1 (0.7) |
| Graft Failure | 1 (0.7) |
| Sinusoidal Obstruction Syndrome | 1 (0.7) |
| Intracerebral Hemorrhage | 1 (0.7) |
| Unknown | 3 (2) |

**Death due to infection in the presence of GVHD is classified as GVHD*

**Table S5.** Organ Involvement by Clinical Grade and Manhattan Score

| Organ Involvement | Grade | Manhattan | n (%) |
|---|---|---|---|
| Isolated UGI | II | Low | 1 (0.4) |
| Skin Stage 1 + UGI | II | Low | 2 (0.7) |
| Skin Stage 3 | II | Intermediate | 42 (15) |
| Skin Stage 2 + UGI Stage 1 | II | Intermediate | 2 (0.7) |
| LGI Stage 1 | II | Intermediate | 41 (15) |
| LGI Stage 2 | III | Intermediate | 27 (10) |
| LGI Stage 3 | III | High | 59 (21) |
| LGI Stage 4 | IV | High | 39 (14) |
| Liver Stage 1 | I | High | 18 (6) |
| Liver Stage 2/3 | III | High | 19 (7) |
| Liver Stage 4 | IV | High | 4 (1) |
| LGI stage 2 + Skin stage 1, 2, 3, or 4 | III | High | 13 (5) |
| Skin Stage 4 | IV | High | 11 (4) |

**UGI, Upper Gastrointestinal; LGI, Lower Gastrointestinal*

**Table S6.** Second Line Treatment by Year

| Years | Ruxolitinib n (%) | Other n (%) |
|---|---|---|
| 2016-2019 | 38 (30) | 87 (70) |
| 2020-2025 | 109 (71) | 44 (29) |

**Figure S1.** Consort Diagram

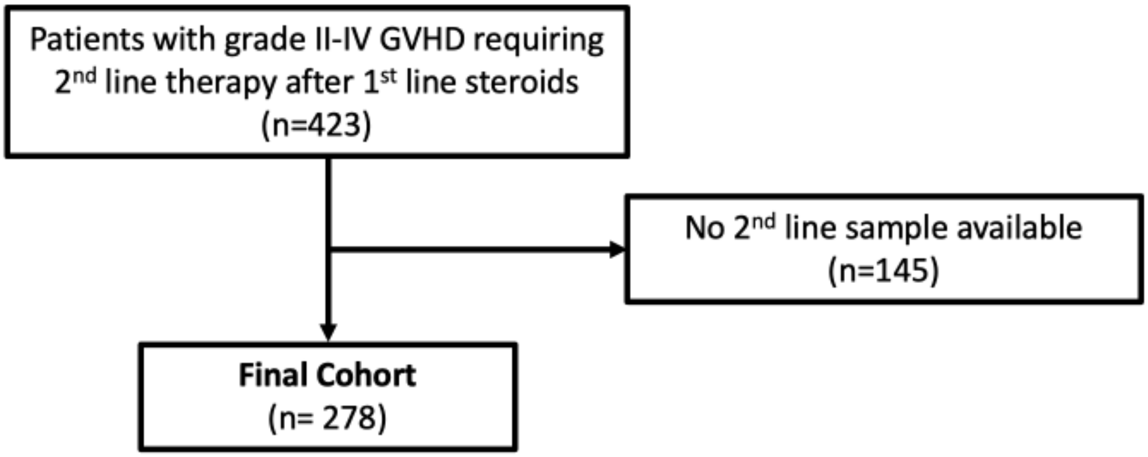

**Figure S2.** Classification by Risk System

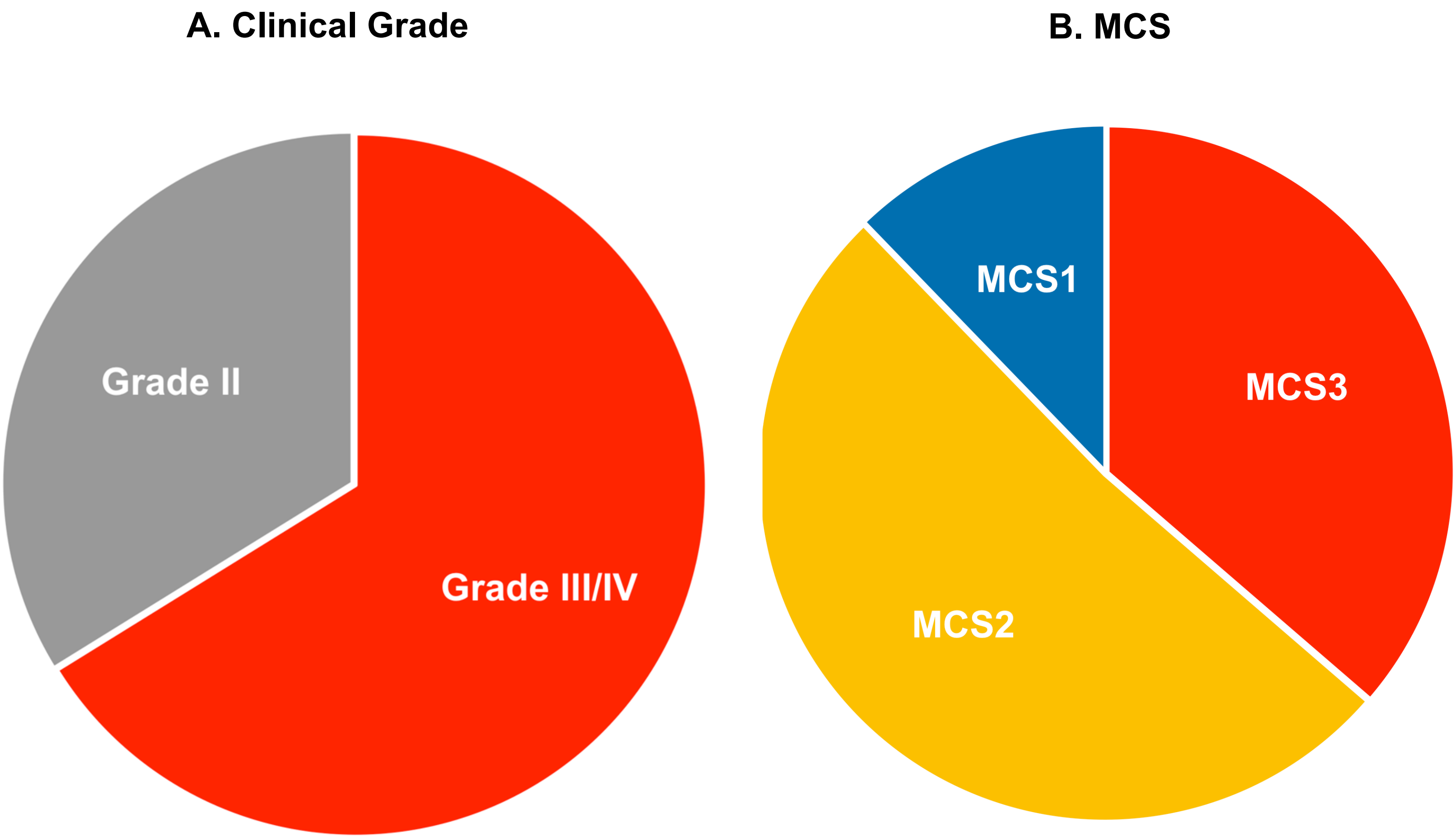

Proportion of patients classified by (A) clinical grade and (B) MCS. 34% (94/278) were classified as clinical grade II and 66% (184/278) as grade III/IV. With Magic Composite, 12% (34/278) are classified as MCS1, 51% (143/278) as MCS2, and 37% (101/278) as MCS3. Integration of biomarkers created an intermediate group (MCS2) that constituted 51% of patients.

**Figure S3.** NRM by Risk System

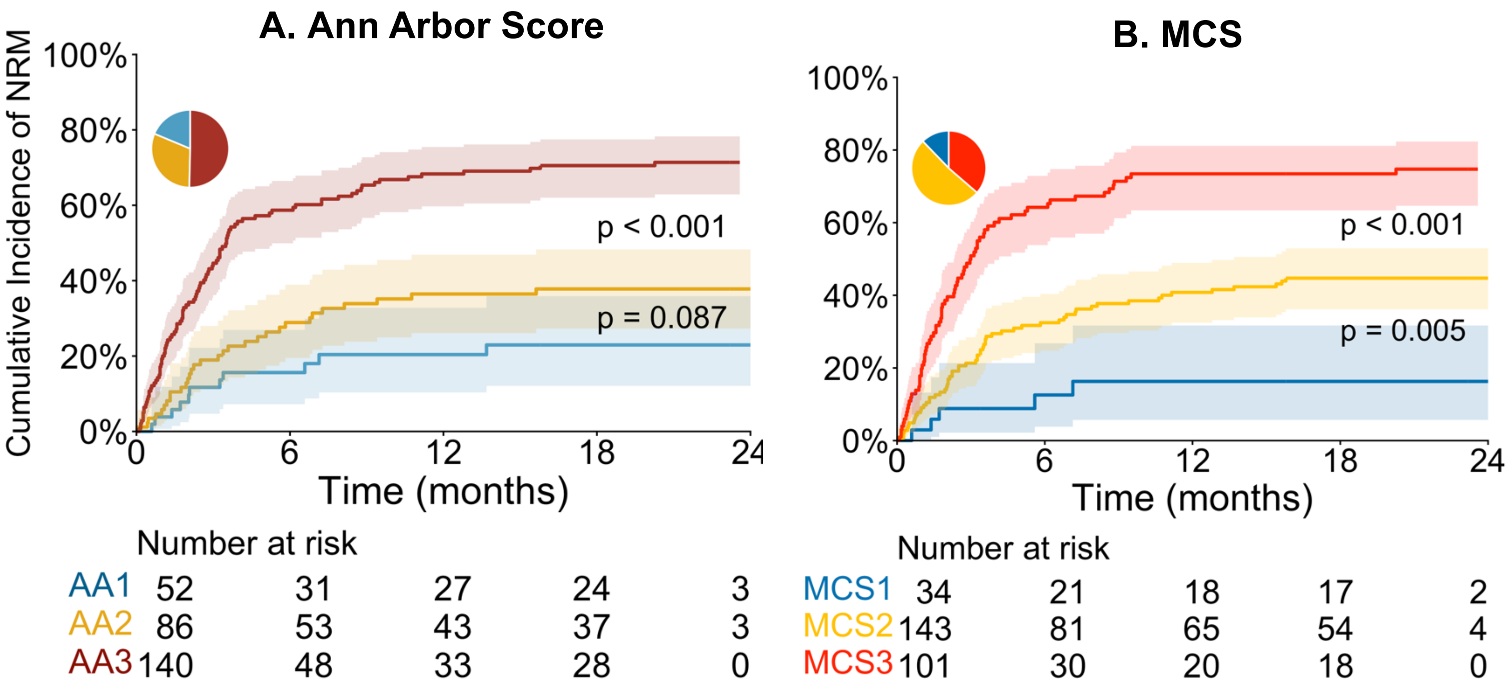

Cumulative incidence of NRM stratified by (A) AA score and (B) MCS. Pie chart depicts the percentage of patients in each risk category.

**Figure S4.** Relapse by Risk System

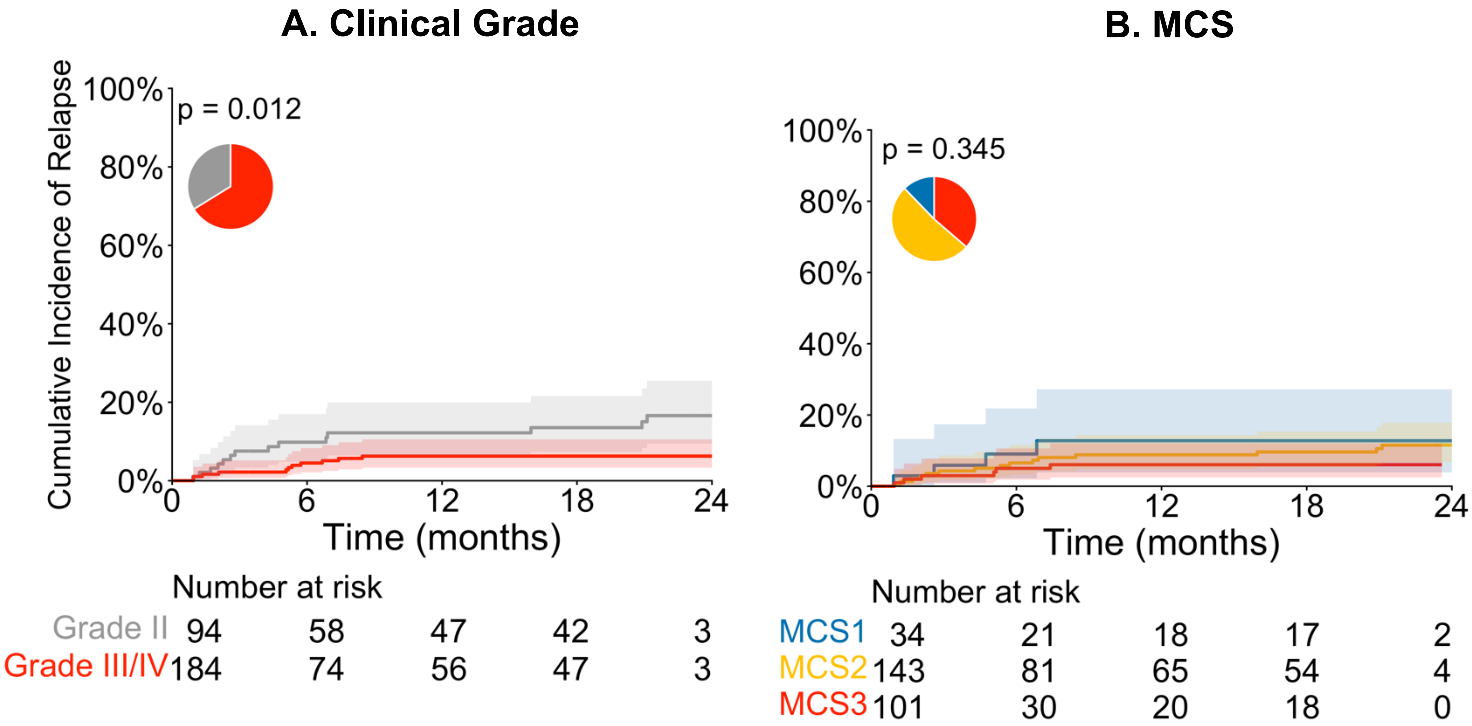

Cumulative incidence of relapse stratified by (A) clinical grade and (B) MCS. Pie chart depicts the percentage of patients in each risk category.

**Figure S5.** NRM and Survival with 2nd Line After 3 Days

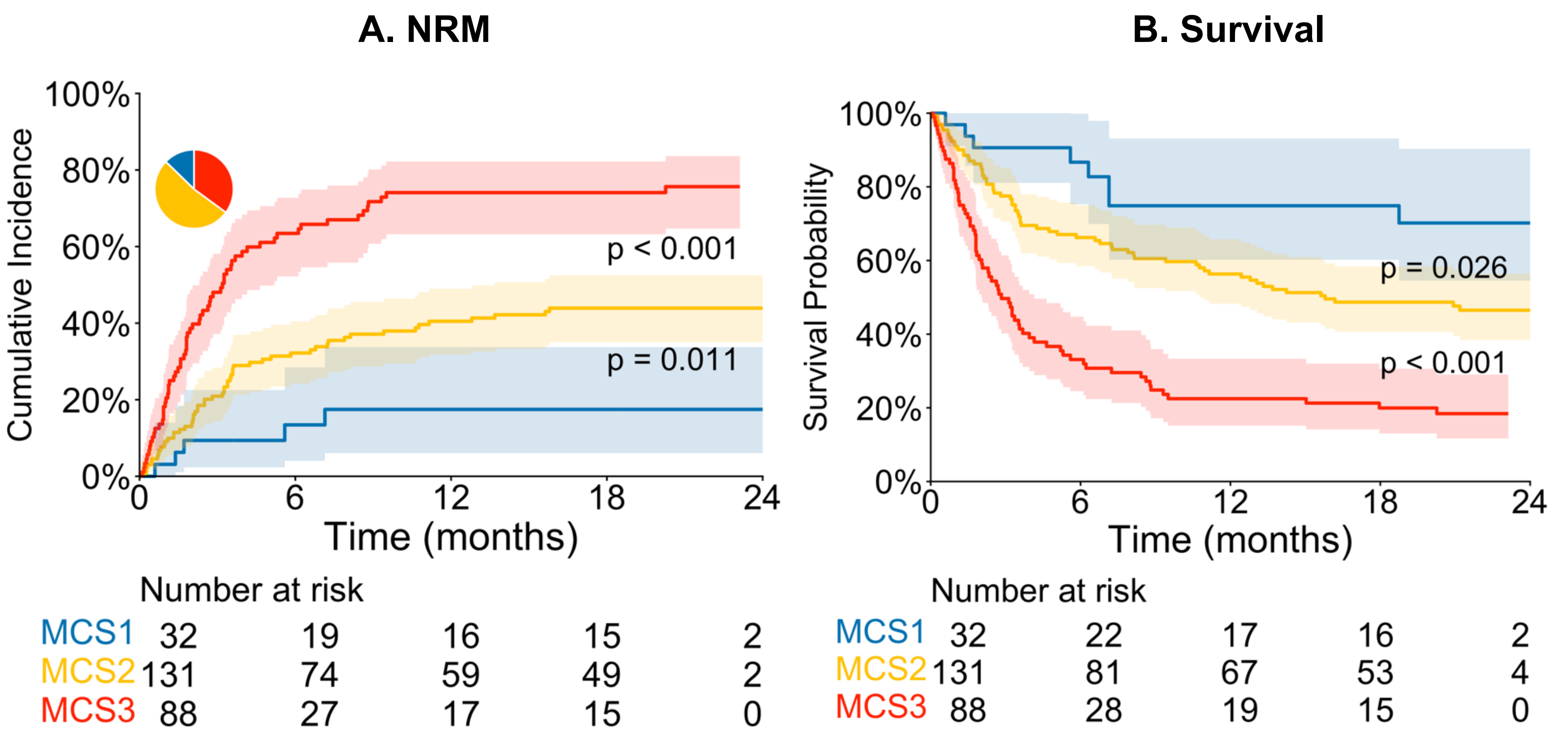

(A) Cumulative incidence of NRM and (B) probability of survival after second-line therapy administered more than three days after first line therapy.

**Figure S6.** Long Term Outcomes by Response to Second Line Therapy

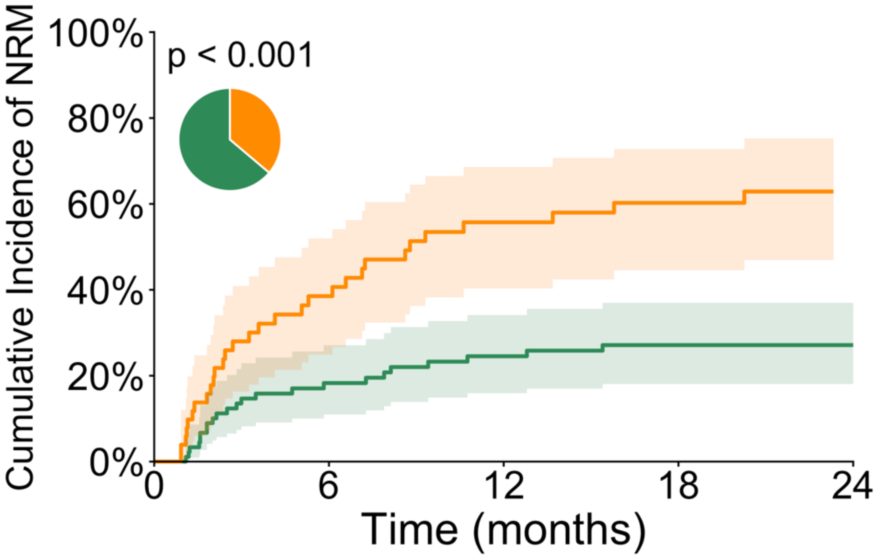

Number at risk

| | | | | | |
|---|---|---|---|---|---|
| CR | 90 | 59 | 49 | 46 | 4 |
| PR | 51 | 26 | 16 | 12 | 0 |

Cumulative incidence of NRM stratified by day 28 response to 2nd line therapy.

**Figure S7.** Day 28 Overall Response by Risk System.

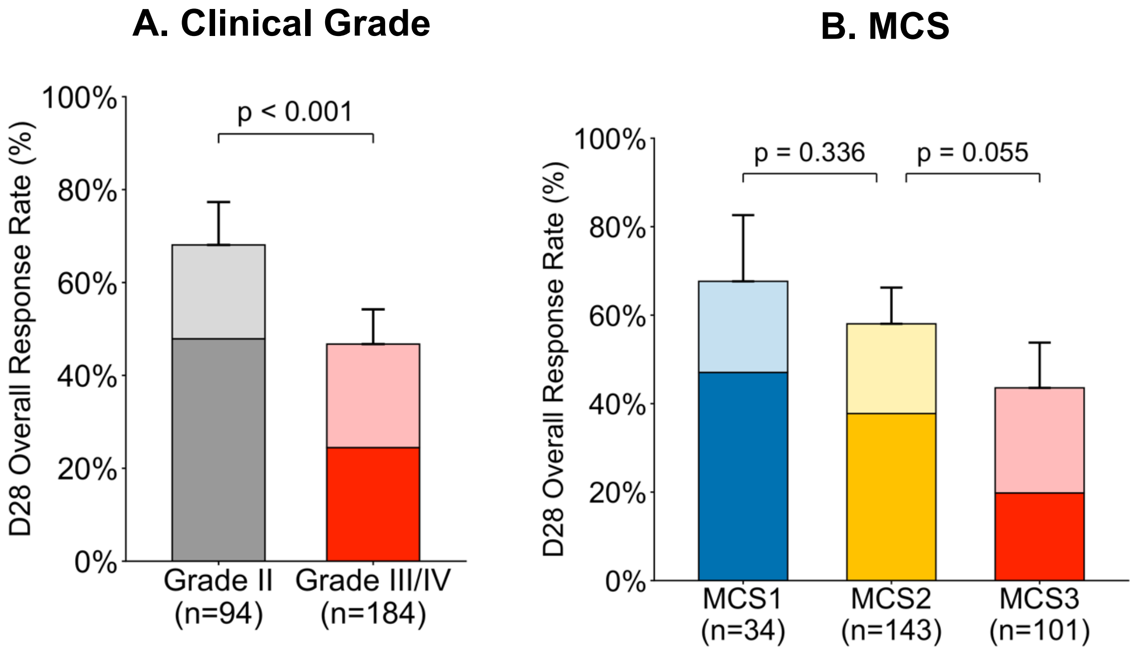

D28 OR rate to 2nd line therapy according to (A) clinical grade and (B) MCS. Lighter shades represent partial response and darker shades are complete response.

**Figure S8.** Long Term Outcomes by 2nd Line Treatment

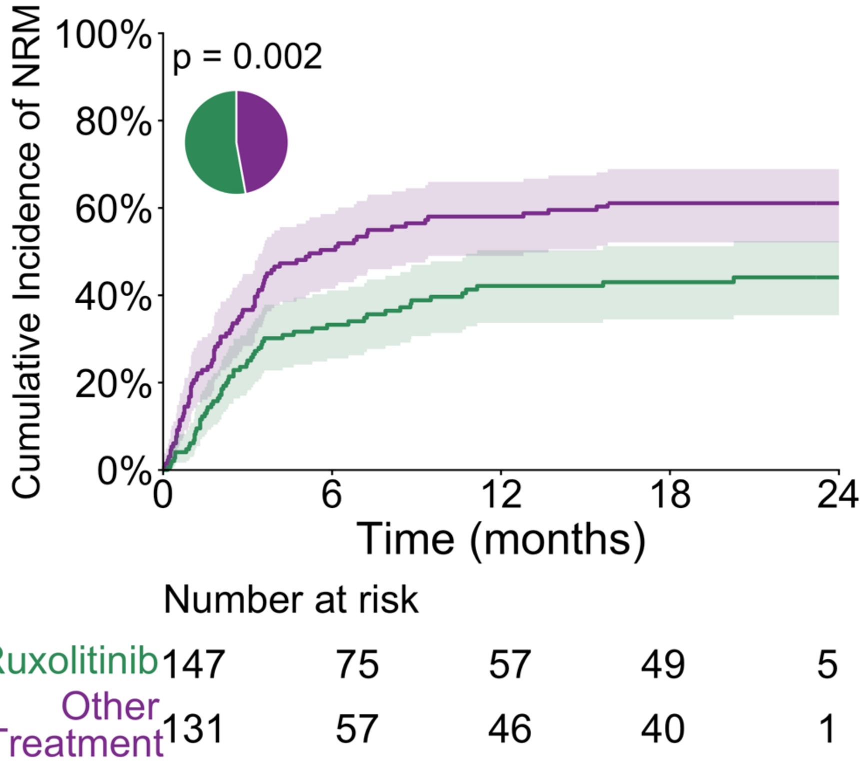

Cumulative incidence of NRM for patients who received ruxolitinib vs other therapies.

**Figure S9**. Day 28 Overall Response to Ruxolitinib by Risk System

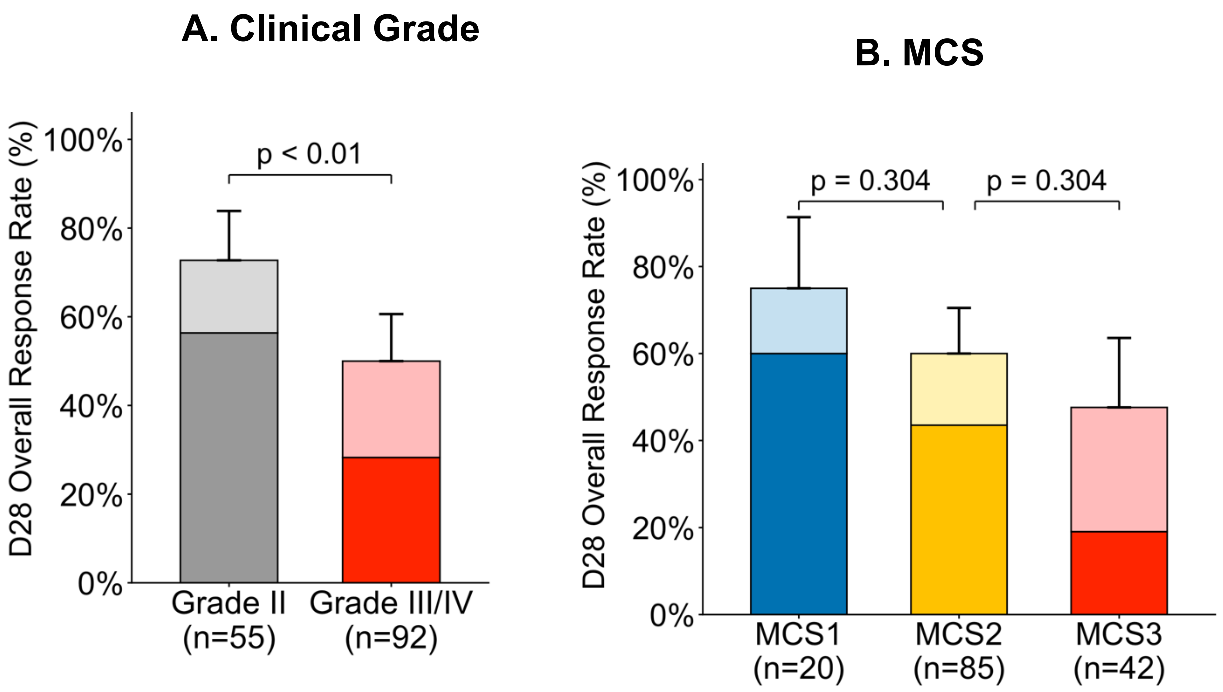

D28 OR rate for ruxolitinib according to (A) clinical grade and (B) MCS. Lighter shades represent partial response and darker shades are complete response.
